# The clinician-AI interface: intended use and explainability in FDA-cleared AI devices for medical image interpretation

**DOI:** 10.1101/2023.11.28.23299132

**Authors:** Stephanie L. McNamara, Paul H. Yi, William Lotter

## Abstract

As applications of AI in medicine continue to expand, there is an increasing focus on integration into clinical practice. An underappreciated aspect of clinical translation is where the AI fits into the clinical workflow, and in turn, the outputs generated by the AI to facilitate clinician interaction in this workflow. For instance, in the canonical use case of AI for medical image interpretation, the AI could prioritize cases before clinician review or even autonomously interpret the images without clinician review. A related aspect is explainability – does the AI generate outputs to help explain its predictions to clinicians? While many clinical AI workflows and explainability techniques have been proposed, a summative assessment of the current scope in clinical practice is lacking. Here, we evaluate the current state of FDA-cleared AI devices for medical image interpretation assistance in terms of intended clinical use, outputs generated, and types of explainability offered. We create a curated database focused on these aspects of the clinician-AI interface, where we find a high frequency of “triage” devices, notable variability in output characteristics across products, and often limited explainability of AI predictions. Altogether, we aim to increase transparency of the current landscape of the clinician-AI interface and highlight the need to rigorously assess which strategies ultimately lead to the best clinical outcomes.

## Main

Applications of AI in medicine are increasingly moving beyond development to clinical integration, especially in imaging domains like radiology. A critical aspect of this integration is where the AI fits in the clinical workflow and the outputs generated to support this workflow. Along with conveying the core prediction of the AI model, these outputs may facilitate explainability in helping the clinician understand how the model arrived at the prediction – a commonly emphasized component for enhancing trust and decision making^1–4^. While many workflow strategies and explainability techniques have been proposed for AI in medical imaging^5,6^, the current scope in clinically-available AI products is not well understood.

To study the current state of the clinician-AI interface, we created a curated database of FDA-cleared AI devices for medical image interpretation, a canonical task among the first to be clinically operationalized. We specifically focus on AI devices with use cases that are historically referred to as variations of “CAD”, a term that stems from computer-assisted detection^7^. As detailed below, there are now several types of CAD that differ according to how the device is intended to be used by clinicians. To create the database, we first identified the FDA Product Codes that support CAD devices. We then reviewed all of the Summary Statements for products with these product codes (see Methods) and curated relevant data, including the intended use and device outputs (see Methods). The final database can be found in the **Supplementary Information**.

We identified 140 FDA clearances from January 2016 to October 2023 for 104 unique AI-enabled CAD products, with some products having multiple clearances over time. The products fall into one of five categories based on their intended use in a clinical workflow, as illustrated in **Figure 1**. These five CAD types vary by their outputs and how clinicians are instructed to use these outputs. For instance, computer-assisted triage (CADt) devices are designed to flag suspicious cases for prioritized review by clinicians. The core AI output for such devices is a binary indicator of whether the case is flagged or not, where flagged cases can be reviewed more quickly by a clinician. Importantly, CADt devices do not provide annotations to directly localize findings^8^. Conversely, computer-assisted detection (CADe) devices help detect the location of lesions by overlaying markings on images. If a numerical or categorical score is assigned to the detected lesion or the whole case, the device is then considered a computer-assisted detection and diagnosis (CADe/x) device because the additional granularity is thought to aid in diagnosis and not just detection^9^. A device that focuses on diagnosis without explicitly marking the locations of lesions across the case is considered CADx. As opposed to CADt devices that flag cases before clinician review, CADe, CADx, and CADe/x devices are designed to assist clinicians as they are interpreting exams. Finally, a variation of CADx has emerged where the device is intended to *automatically* interpret the exam without clinician review^10^. We denote this use as CADa, which is currently only used for one specific application as discussed below.

**Figure 1.**
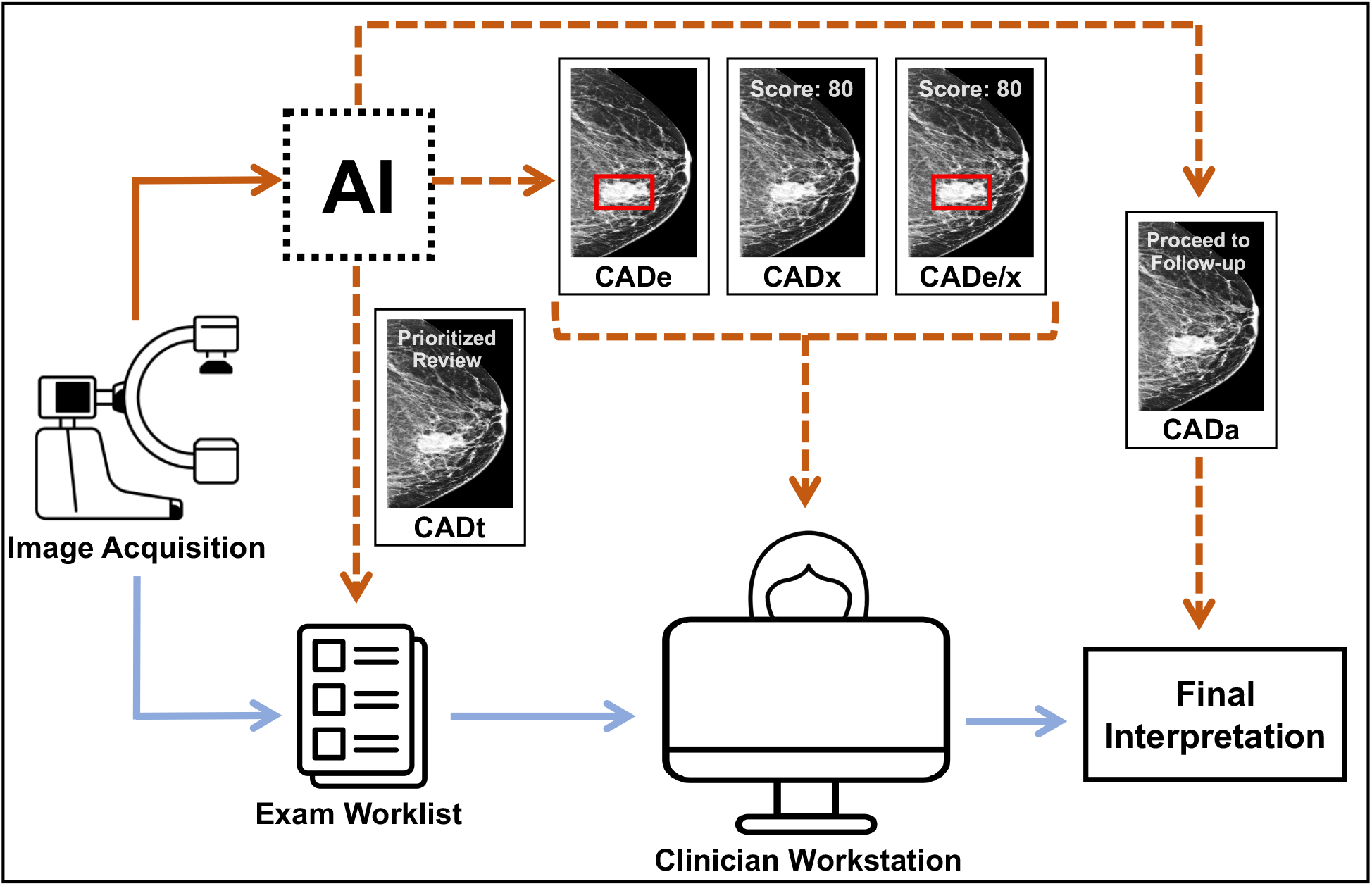
Overview of types of FDA-cleared CAD products and their integration into medical image interpretation workflows. CAD types vary according to their outputs and place within the clinical workflow. CADt (triage) devices are designed to flag cases for prioritized review and do not place marks on the image. CADe (detection) devices mark regions of interest to aid in the detection of lesions as a clinician is interpreting an exam. CADx (diagnosis) devices are designed to aid in diagnosis, such as by outputting a score or category, but do not explicitly detect lesions across the exam. CADe/x (detection & diagnosis) devices provide both detection and diagnosis support. Finally, an autonomous system, which we denote as CADa, aims to automatically interpret the exam without clinician input.

A breakdown of the CAD types across the 104 FDA-cleared products is shown in **Figure 2a**. CADt is the most frequently approved product type, representing 59% of products, followed by CADe with 19%. The distribution of CAD types is also highly dependent on the disease, with some diseases having multiple CAD types and others only one (**Figure 2b**). Breast cancer and intracranial hemorrhage (ICH) have the highest number of products with 14 each. CADt, CADe, CADx, and CADe/x are all represented in breast cancer, whereas all of the ICH products are considered CADt. Altogether, we find 37 different diseases/conditions represented, with conditions with more than three products shown in **Figure 2b**.

**Figure 2.**
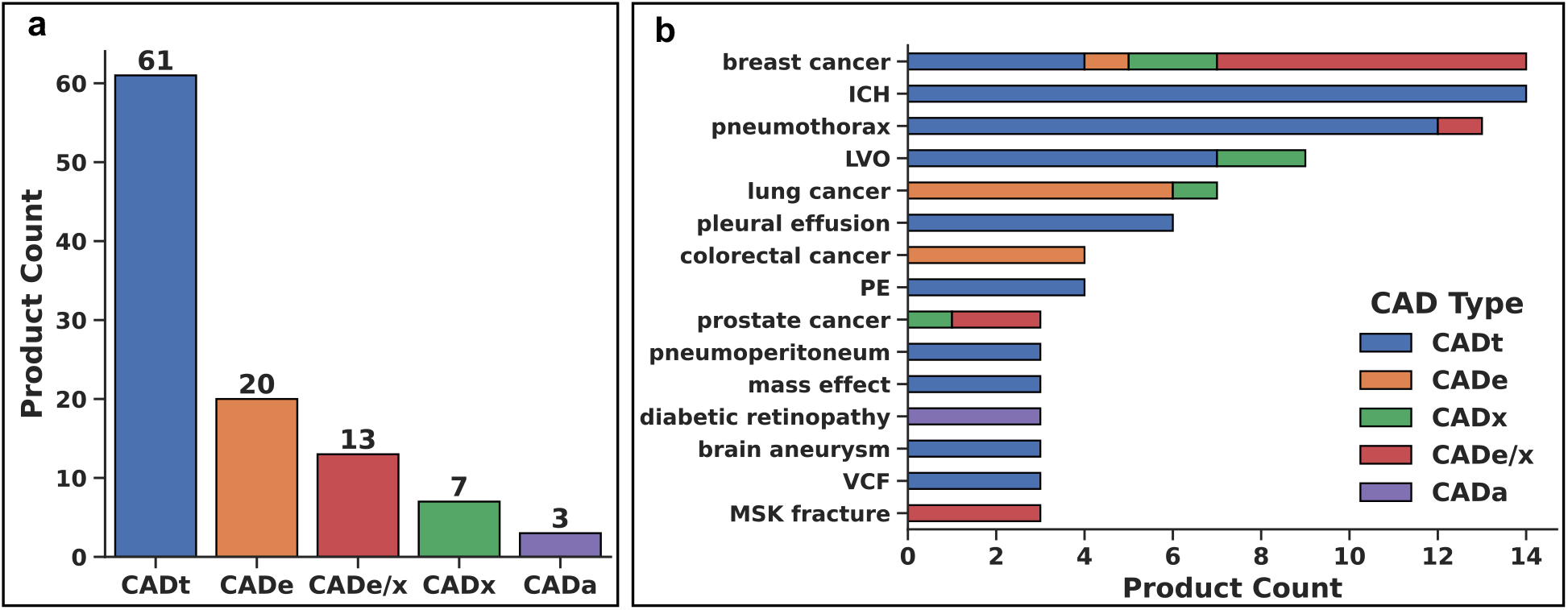
Landscape of intended uses of FDA-cleared AI products for medical image interpretation. a) Total number of FDA-cleared AI products from January 2016 to October 2023 for each CAD type: CADt (triage), CADe (detection), CADx (diagnosis), CADe/x (detection & diagnosis), CADa (autonomous). b) Distribution of FDA-cleared AI products for each CAD type by disease indication. Diseases/conditions with three or more products are shown. ICH: intracranial hemorrhage; LVO: large vessel occlusion; PE: pulmonary embolism; VCF: vertebral compression fracture; MSK: musculoskeletal.

Beyond CAD type, we curated finer details regarding the outputs of FDA-cleared AI devices. From a practical standpoint, we can consider these outputs to have two functions: 1) convey the core prediction of the AI model to help with the final diagnosis, and 2) convey information to support this prediction. For instance, an AI model may predict that a head computed tomography (CT) exam is suspicious for ICH (the core prediction) and also indicate the location of the hemorrhage or show similar examples from the training dataset where ICH was also present. These additional outputs can be considered a form of explainability in facilitating clinician understanding and trust of the prediction.

Across the database, we find high variation in output characteristics of the AI devices. This variation is present both in terms of the form of the core prediction and the presence and type of explainability. Starting with the core prediction, we categorized each product as having a binary, categorical, or score-based prediction output. For example, a product may characterize an exam/lesion as suspicious or not (binary), low vs. medium vs. high suspicion (categorical), or generate a suspicion score between 1-10 (score-based). **Figure 3a** illustrates the distribution of prediction output types across the AI products. We find that binary-level predictions are by far the most common across FDA-cleared products. This is in large part driven by CADt and CADe products that generate binary-level predictions at the case- or lesion-level, respectively. Categorical and score-based outputs are nonetheless represented in CADx and CADe/x products, though categorical outputs are three-times less common than numerical scores.

**Figure 3.**
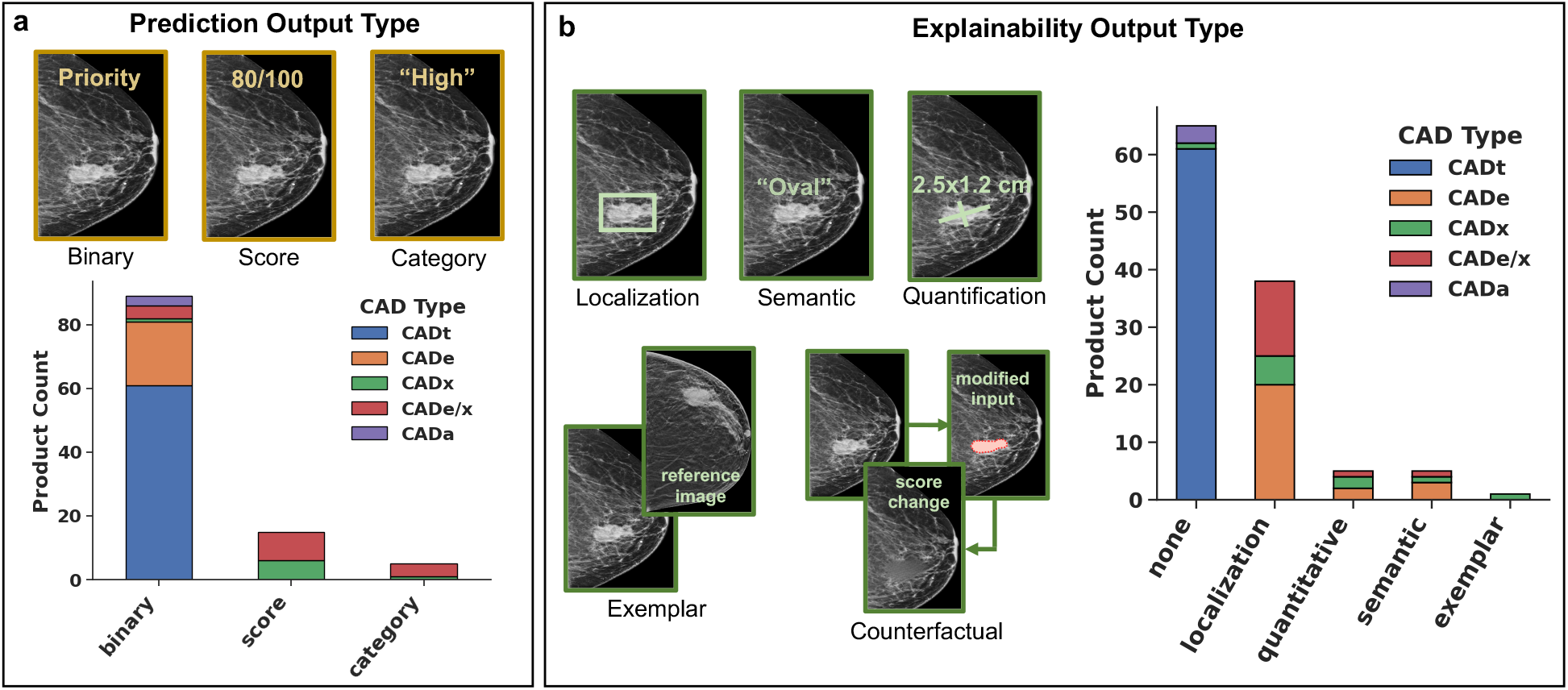
Prediction and explainability output types of current FDA-cleared AI products for medical image interpretation. a) Predictions are grouped according to binary, category, or score. b) Type of explainability offered by products, with “none” corresponding to products that provide image/exam-level predictions without explicit localization or other form of explainability. Counts are also indicated by CAD type: CADt (triage), CADe (detection), CADx (diagnosis), CADe/x (detection & diagnosis), CADa (autonomous).

Beyond prediction type, we curated the type of explainability offered by the AI products, considering several types of explainability that have been discussed in AI literature^5^. We consider explainability from a user interface perspective and group product outputs according to several categories that are illustrated in **Figure 3b**. Localization-based explainability can take different forms such as bounding boxes or heatmaps, where these outputs help convey the “where” behind an AI model’s prediction. Other types of explainability also convey aspects of “why” or “what”. For instance, an exemplar-based explanation might retrieve and display reference examples in the training dataset that have similar qualities to the image under consideration. An approach that is becoming increasingly popular in AI research is the use of counterfactual explanations^11^ and related generative techniques. A counterfactual approach involves minimally modifying the image to flip an AI model’s prediction, thus giving intuition on the features used by the model in making the original prediction. Other explainability categories include the use of language-based semantics or quantitative characteristics. For instance, an AI model may characterize a detected lesion as “round” or estimate its size as 2 cm, both of which may help the clinician understand and trust (or be skeptical of) a model’s prediction.

The distribution of explainability types in the FDA-cleared AI devices is illustrated in **Figure 3b**, where “none” corresponds to image/case-level predictions without explicit localization or other types of explainability. Although we do not find examples of counterfactual explanations, each of the other described categories of explainability are represented across the products. Not surprisingly, “none” is the most common category of explainability given the popularity of CADt, which does not offer explicit localization or other explainability types. When a form of explainability is provided, localization is by far the most common, followed by semantics, quantitative, and exemplar with 5, 5, and 1 products, respectively.

In summary, while several studies have analyzed aspects of FDA-cleared AI devices^12–14^, there is a pressing need for enhanced transparency around factors related to clinical integration. To this end, we assembled and analyzed a curated database focusing on the canonical use case of medical image interpretation assistance (“CAD”). Our analysis finds 140 FDA clearances for 104 products across five different CAD types. By far the most frequent CAD type is CADt, where there are more products with this triage use case than all other types combined. While CADt products are constrained in the types of outputs provided, we find meaningful variation in core user interface parameters for products of other CAD types. Nonetheless, usage patterns are highly skewed, with score-based predictions more popular than categorical, and localization-based explainability being the most common technique when a form of explainability is offered.

The optimal AI-clinician integration strategy depends on a number of factors, yet even seemingly minor differences in AI outputs may ultimately lead to dramatic differences in clinical efficacy. As providers consider AI adoption, it is especially instructive to be aware of the different CAD types and their advantages and limitations across different diseases. In the case of CADt, several studies have indeed shown the potential for faster turnaround times and improved outcomes for prioritized exams^15,16^. However, the FDA has also recently released a letter “reminding health care providers about the intended use of radiological computer-aided triage and notification (CADt) devices for intracranial large vessel occlusion (LVO)”, including statements that such devices are not diagnostic and cannot rule out the presence of an LVO^17,18^. As such, the AI output for CADt devices is minimal and the core study required for FDA clearance is standalone AI performance testing^8^. Appreciating clinician-AI considerations is similarly important for AI developers in envisioning how a core AI model could fit into existing or new workflows and aligning model development with this in mind. There are especially opportunities to assess whether recently popular explainability techniques such as counterfactual and text-based explanations can improve clinical utility, as these techniques are not yet robustly represented in current products. Altogether, rigorously studying the clinician-AI interface will help accelerate the clinical translation of AI in a safe and effective manner.

## Methods

To curate a list of FDA-cleared AI CAD products, we first identified the FDA Product Codes that support CAD devices by reviewing all Product Code descriptions^19^ in both a manual and keyword-search manner, resulting in the following list: MYN, OEB, PIB, POK, QAS, QBS, QDQ, QFM, QNP, QPN. We note that other product codes that may include forms of image processing but are not explicitly indicated for CAD-based assistance, such as LLZ and QIH, were not considered. From the final list of Product Codes, we then retrieved a list of all products for these codes using the FDA’s 510(k)^20^, De-Novo^21^, and PMA^22^ databases. From the summary statement for each product, we manually extracted the intended use, device outputs and inputs, and types of algorithms used. Each product was independently reviewed and confirmed by two researchers. For a small number of products where the summary statement was ambiguous, we additionally consulted online product documentation. As our goal was to study products based on modern deep learning-based AI techniques, we excluded any products that describe the use of purely traditional (shallow) machine learning or hand-engineered computer vision techniques. We additionally compared our final product list to a list released by the FDA that covers AI-enabled products with clearances through July 2023^23^ to ensure consistency across the overlapping time period for our identified product codes. Finally, we cleaned and standardized the extracted data to maintain standard nomenclature across products. We additionally identified which clearances are new versions of previous products versus new products altogether, which we determined based on a product having a consistent name/manufacturer, intended use, and disease indication(s) as a prior clearance. The final curated database can be found in the **Supplementary Information**.

We characterized the outputs of each product according to the form of its core prediction and type of explainability offered. As each device is indicated for a specific disease(s)/condition(s) we consider the core prediction to be the device’s estimate of the presence of this disease(s)/condition(s). This prediction could take a number of forms, which we grouped into three buckets: binary, categorical, or score-based. Categorical predictions consist of text-based classifications such as “high” vs. “medium” vs. “low”. Score-based predictions consist of numerical outputs, typically with at least 10 increments (e.g., 1-10). In circumstances where a prediction has aspects of both a text identifier and number (generally with <10 increments), the prediction was considered categorical. An example of this would be the BI-RADS classification system which consists of a number from 0-6 and a text category corresponding to this number. Additionally, a given product may have more than one output type, such as both categorical and score-based predictions, in which case each type was included in the final tally. If a product had more than one clearance, such as an update over time, the most recent version was used for all analysis.

## Data Availability Statement

The curated database used for all analysis is available as Supplementary Material.

## Supporting information

AI Device Database

